# PROLONGED LENGTH OF HOSPITAL STAY AND ASSOCIATED FACTORS AMONG PATIENTS ADMITTED AT A SURGICAL WARD IN SELECTED PUBLIC HOSPITALS ARSI ZONE, OROMIA, ETHIOPIA, 2022

**DOI:** 10.1101/2022.10.18.22281234

**Authors:** Deriba Fetene, Yohanis Tekalegn, Jabir Abdela, Amdehiwot Aynalem, Gezahegn Bekele, Ezedin Molla

## Abstract

**Background:** Prolonged length of hospital stay is the most common indicator of poor quality of health care and inefficient utilization of hospital resources. Prolonged hospital stay associated with increased mortality, hospital-acquired infection, and unnecessary utilization of hospital bed and other resources. Predicting length of hospital stay facilitates resource designing and initiates quality improvement activities. However, data regarding the prolonged length of hospital stays, and associated factors were a scarce in Ethiopia.

**Methods:** A hospital-based cross-sectional study was conducted on a sample of 316 adult patients from December 1 to January 10, 2022. A consecutive sampling technique was used during sampling procedure. A structured questionnaire was used to collect data regarding sociodemographic factors, clinical factors, medication factors, and behavioral factors through interview, medical record review and by using check list. The data was entered into the epidata4.6 version and exported to SPSS Version25 for binary logistic regression analysis. To identify factors associated with outcome variable, candidate variables (p<0.25) were fitted to multivariable analysis, and those with P-values<0.05 were considered as factors associated with prolonged length of hospital stay. Odds ratio with corresponding 95% CI was used to indicate the strength of association of variables with prolonged length of hospital stay.

**Result:** One fourth (24.7%) of the study participants experienced a prolonged length of stay at surgical ward. The odd of a prolonged length of hospital stay was high among patients who had comorbid condition [AOR=4.59, at 95% CI= (2.46-8.56)], who developed surgical site infection [AOR=5.02 at 95% CI= 1.97-12.80)], and who developed hospital acquired pneumonia during hospital stay [AOR= 3.43 at 95% CI= (1.36-8.64)].

**Conclusion and recommendation:** Near to quarter of the study participants’ experienced prolonged length of hospital stays. Comorbid condition, surgical site infection, and hospital acquired pneumonia were factors associated with prolonged length of hospital stay at surgical ward. Quality of care could be improved by adjusting surgical ward environment to prevent hospital acquired infection and focus on managing complication after surgery. Health care provider should be educating surgical patient about the risk of comorbidity on wound healing and early diagnosis and prevention of comorbid condition.

## 1. Introduction

Hospital length of Stay (LOS) is one of the most common indicators to measure the quality of health care service and efficient utilization, allocation and administration of hospital resources. It refers to the total bed days occupied by patient from the time of admission until discharge (Marfil-Garza et al., 2018). Prolonged length of hospital stay (PLOS) which is defined as hospital stay more than expected length of stay (LOS) for a certain procedure resulted in hospital acquired infection, mortality, inessential utilization of hospital bed, and capacity shortage (Lee et al., 2018).

Hospital length of Stay (LOS) is operationalized as 75^th^ percentile length of stay, and by this measure patient’s length of stay classified as those with normal length of stay (NLOS) and those with a prolonged length of stay (PLOS). The study in Singapore revealed that below median LOS as normal and above median (50^th^ percentile) cutoff point with length of stay for at least 6 days as a prolonged length of stay (PLOS) (Lee et al., 2018). In Harvard medical school prolonged length of hospital stay was defined as length of stay above 75^th^ percentile (at least 8 days) (Dasenbrock et al., 2015). Also, other studies were used 75^th^ percentile cutoff point to determine a prolonged length of stay. This studies include study conducted at Michigan university in America with length of stay for at least 9 days (Krell et al., 2014), at Oman tertiary hospital in Iran with length of stay for at least 11 days (Almashrafi et al., 2016), and study done at Jimma University medical college (JUMC) in Ethiopia with length of stay for at least 33 days(Tefera et al., 2020). 95^th^ percentile is another definition used as NLOS and PLOS, in Mexico prolonged length of stay (PLOS) was considered as length of stay above 95^th^ percentiles (at least 34 days) and below 34 days as NLOS (Marfil-Garza et al., 2018).

Prolonged length of hospital stays (PLOS) after cardiac surgery can cause excessive utilization of hospital resources. In Iran 30.5% of the patients who undergo cardiac surgery experienced PLOS (Almashrafi et al., 2016). Even though the length of hospital stay is one of the tools used to measure quality of care and efficient utilization of hospital resources, patients with PLOS were exposed to different problem. Length of hospital stay (LOS) among patients admitted at surgical unit has been decreasing in developed countries but, in Africa it is still high despite its unwanted effect on patients and hospital. In Nigeria Ibadan hospital PLOS has been shown to be a source of embarrassment to family members. Unable to carrying out different investigation after surgery, buying of post-operative medications and payment of hospital bill is progressive among patients who spent longer time in Nigeria Ibadan hospital (Ilesanmi and Fatiregun, 2014). Patients with under nutrition had extended hospital stay because of decrease wound healing process, increase hospital complication, and burden of health care cost. Previous studies identified that 40% of patients are under nutrition during admission and these patients continued to be malnutrion throughout hospital stay. Patient with malnutrition status have death and hospital complication 3-4 times higher than well-nourished patients with PLOS (Abrha et al., 2019).

Prolonged length of stay could cause multiple effects on patients and hospital. Among this surgical site infection and hospital acquired pneumonia, delay of critically ill patients timely access to treatment, increase mortality rate by threshold, and high expenditure for health cost (Lee et al., 2018). Unnecessary utilization of hospital bed and medication that result in capacity shortage are some of problem associated with PLOS (Tefera et al., 2020).

No study have been conducted regarding prolonged length of stay and associated factors among patients admitted in surgical unit at Asella teaching and referral Hospital (ATRH) and Bekoji District hospital (BDH). In Ethiopia, little is known about PLOS and associated factors, in order that this study is aimed to assess the magnitude of a PLOS and associated factors among adult patients admitted at surgical ward of ATRH and BDH.

This study finding will be important in minimizing hospital-acquired pneumonia and surgical site infection. It also avoid delay in elective surgery due to unnecessary occupy of surgical bed, facilitating timely access to treatment, and decreasing economic burden of patients and family. Additionally, knowing the factors that increase LOS in surgical unit can allow hospital staff to manage the delivery of quality of care and demand for bed after the patients admitted to the ward that increase effectiveness in utilization of hospital resources. By identifying the major problem associated with the prolonged length of hospital stay (PLOS) in the study area, the study will also help to inform policy maker to implement specific and other relevant option to develop efficient program that would benefit public hospitals found in Arsi zone.

## 2. Methods

### 2.1 study area and period

The study was conducted at Arsi Teaching and Referral Hospital (ATRH) and Bekoji district hospital (BDH) in Arsi zone, Oromia region, Ethiopia from December 1 to January 10, 2022. ATRH was previously called Haile Selassie first hospital and established in 1972 E.C for the purpose of health professionals’ training and health care service. Arsi Teaching and Referral Hospital is found in Asella town Oromia region. It has 08 inpatient units with a total capacity of 300 beds serving around 20 million people. Currently, 442 health professionals are working in this hospital, of which 212 were nurses. The surgical ward (SW) has a total of 54 beds and 3600 patients were admitted to this ward per year. Bekoji District Hospital (BDH) which found in Bekoji town was established in 2007 E.C as a primary hospital. It has 08 inpatient units with a total capacity of 182 beds serving around 18 million people. Currently, 400 health professionals are working in this hospital, of which 150 were nurses. The Surgical ward has a total of 20 beds with 2400 admission per year to this ward.

### 2.2 Study design

A hospital-based cross-sectional study was conducted

### 2.3 Population

All patients admitted to the surgical ward of ATRH and BDH during the study period were source population. Selected adult patients admitted to the surgical ward of ATRH and BDH during the study period was taken as study population.

### 2.4 Eligibility criteria

All sampled adult patients admitted to surgical ward at ATRH and BDH with age of ≥18 years old (yo) were included in the study. On the contrary, those patients who were severely ill were excluded from the study.

### 2.5 Sample size and sampling technique

The sample size (n) was calculated by using a single population proportion formula by considering 1.96 for the standard normal variable with a 5% level of significance (α value), 95% confidence interval, 5% margin error, and 25.3% proportion from previously conducted study (Tefera *et al*., 2020). By adding 10% (29) nonresponse rate, the final sample size was 319. Two public hospitals, Arsi teaching and referral hospital and Bekoji district hospital were selected from total of 8 public hospitals identified in Arsi zone using simple random sampling method. To allocate the number of the study participants from SW at ATRH and BDH, a one year SW patients record was assessed and the trend showed that 6000 (3600 from ATRH and 2400 from BDH) were admitted in year 2013 E. C. Based on this, sample size distribution was done as follows: 191 patients [(300/500) ×319] from ATRH and 128 patients [(200/500)×319] from BDH were included in the study. Consecutive sampling technique was used in which every patients meeting the inclusion criteria were selected until the required sample size was achieved.

### 2.6 Dependent variable

Prolonged length of hospital stay is dependent variable

### 2.7 Independent variable

Independent variables are socio-demographic factors (age, sex, marital status, educational status, place of residence, occupation, monthly income, and health care cost coverage), clinical factors (time of admission (local time), comorbid condition, type of surgery, amount of blood loss during surgery, type of anesthesia, waiting time for surgery (in day), surgical site infection, hospital acquired pneumonia), medication factors (medication used in the past 3 months, medication used after admission, number of medication used during hospital stay), behavioral factors (body mass index, smoking status, alcohol status).

### 2.8 Operational definition

Length of hospital stay: Is the duration of a single episode of hospitalization which is calculated by subtracting the day of admission from the day of discharge (Tefera *et al*., 2020) Normal length of hospital stay: Hospital stays less than the 75^th^ percentile length of stay was used (Tefera *et al*., 2020)

Prolonged length of hospital stay: Hospital stays greater than the 75^th^ percentile was used (Tefera *et al*., 2020)

Smoking status: Who smoke in the past 3 months from the time of data collection (Jamal et al., 2018)

Alcohol drinking status: Those who drink alcohol in the past 12 months from the time of data collection (Tyrovolas et al., 2020)

Number of medication: Total number of medication taken by patients during hospital stay and classified as ≤2 and >2.

### 2.9 Data collection tool and procedure

Data collection instrument was adapted from the study done in Jimma University medical center. The tool was prepared in English and translated to Amharic and afaan Oromo version with the assistance of a language expert. Data were collected by a structured and face to face interview, medical record review, and observational checklist. The assessment tool had four parts. The first part was about socio-demographic factors with 08 items. The second part assessed clinical characteristics with 08 items. The third part was medication-related questions with 03 items, and the last part was about behavioral related tool with 03 items. Observational checklist used for assessment of hospital acquired pneumonia and surgical site infection were adapted from world health organization (WHO). Data was collected by two professional and trained nurses and two supervisors. The collected data was checked daily for consistency, accuracy, and preventing any bias.

### 2.10 Data quality control measures

The training for data collectors and the supervisor was given by the principal investigator for two days regarding the approach to the patients and the confidentiality before data collection. The questionnaire was pretested on 5 % (16) of the sample at Arsi robe hospital two weeks before conducting data collection to check acceptability and consistency. Depending on the result of the pretest slight modification of the questionnaire was done for clarity and completeness of the questionnaire. Continuous supervision and daily checking of the collected data were performed by investigator.

### 2.11 Data processing and analysis

Data were coded, entered, and cleaned using Epi-data 4.6 version. Then the data was exported to SPSS (statistical package for social science) version 25 for analysis. Descriptive statistics like frequency and percentage was used to summarize the characteristics of the study participants. Continuous variables were expressed using measures of central tendency and variability such as mean and standard deviation. The binary logistic regression model was used, and bivariate logistic analysis was done to identify candidate variables for multivariate logistic regression at P-values <0.25. Odds ratio with 95% confidence interval (CI) was done to predict the presence and strength of association between outcomes and independent variables. Finally, the level of significance of an association was considered at a p-value of < 0.05 in a multivariate logistic analysis.

## 3. Result

Out of total of adult patients admitted at surgical ward, 316 participated in the study yielding response rate of 98.7%.

### 3.1 Sociodemographic characteristics of participants

The mean (± SD) age of the study participants was 38.31±15.84 years. 216(68.4%) of the study participants were male and 196(62%) were married. 163(51.6%) were living in the rural area and 281(88.9%) of the study participants were paid the cost for health coverage by themselves (see table 1).

**Table 1:**
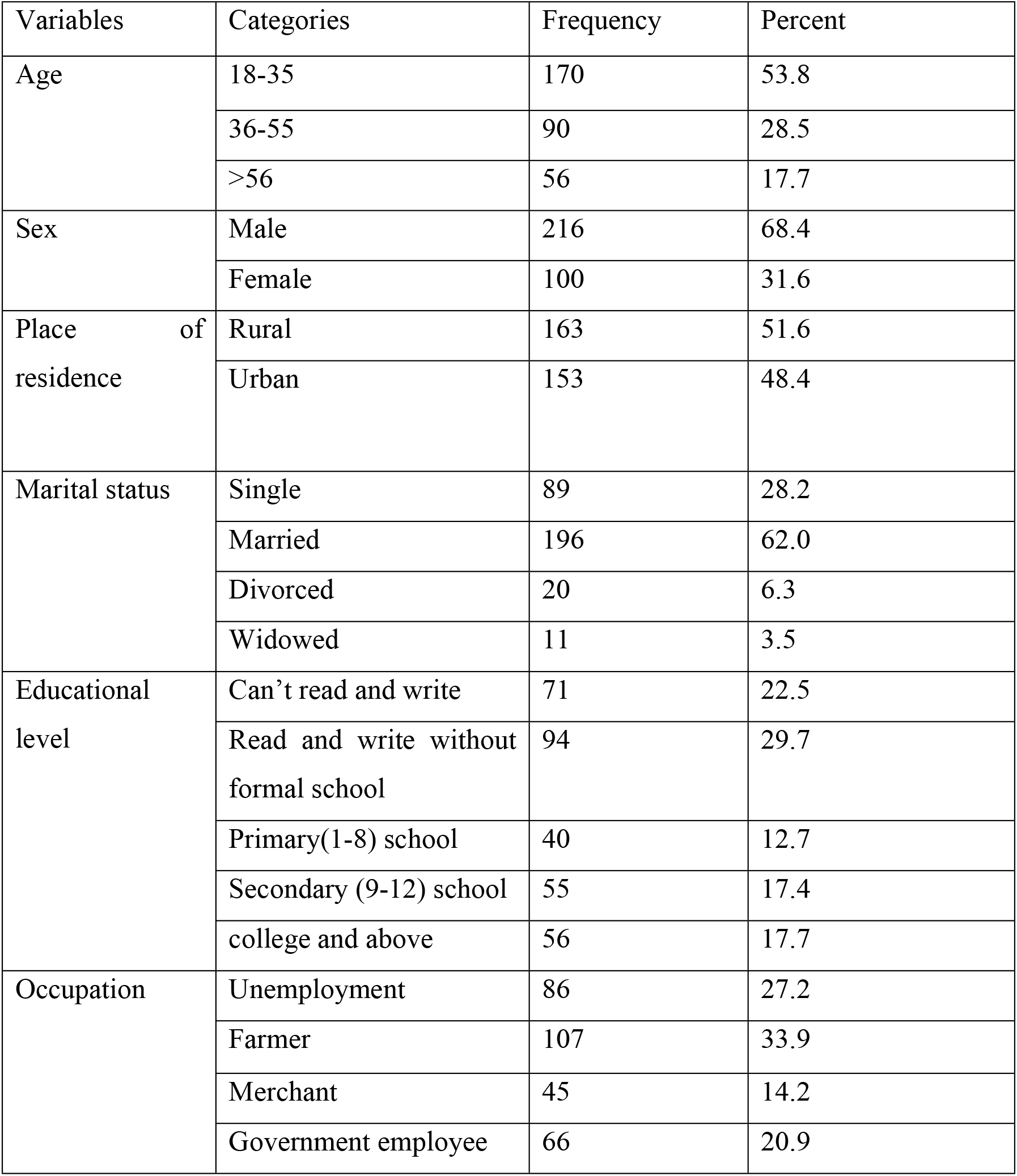
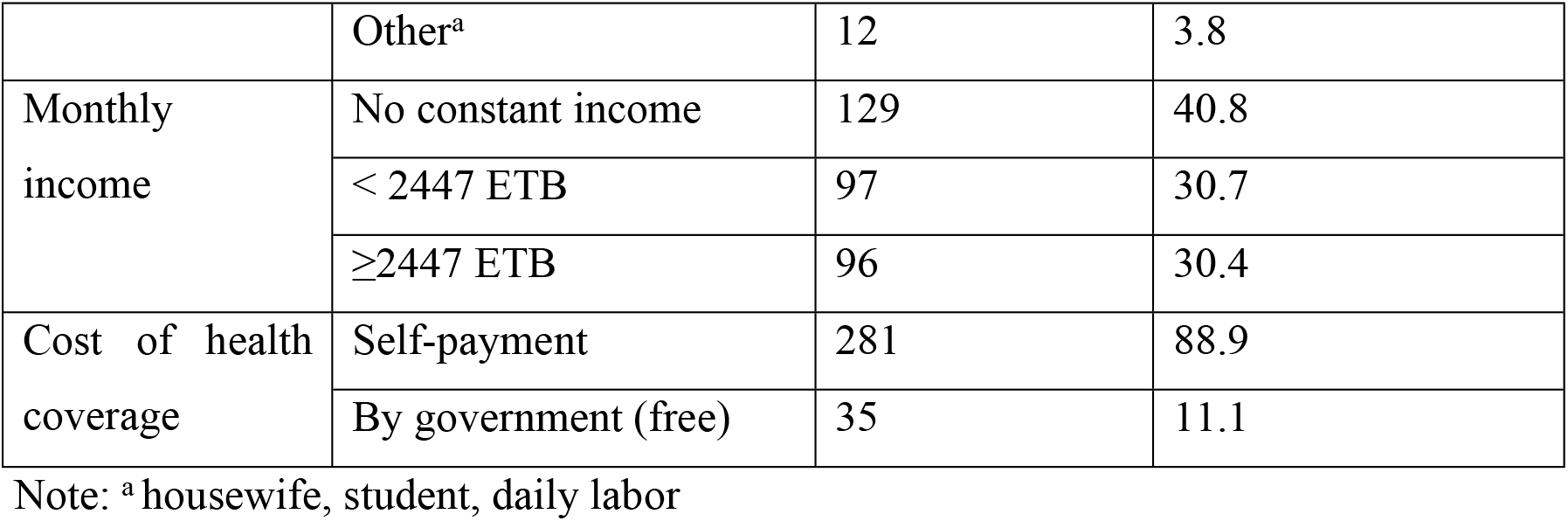
Sociodemographic characteristics of the study participants at surgical ward of ATRH and BDH, Arsi zone, Oromia, Ethiopia, 2022 (n=316)

### 3.2 Clinical characteristics of the participants

Among 316 of the study participants 238(75.3%) were stay in the surgical ward less than 12 days and 78(24.7%) stay above. From 261(82.6%) of the study participants undergo surgery; 151(47.8%) was elective. 81(25.6%) of the study participants had comorbid condition, 30(9.5%) developed surgical site infection, 26(8.2%) experienced hospital acquired pneumonia respectively. 15(4.7%) of the patients have been used medication in the past 3 months and 316(100%) used medication after admition. 145(45.9%) used more than 2 medication during hospital stay (see table 2).

**Table 2:** Clinical characteristics of the study participants at the surgical ward of ATRH and BDH, Arsi Zone, Oromia, Ethiopia, 2022 (n=316)

| Variables | Categories | Frequency | percent |
| --- | --- | --- | --- |
| Length of stay | Less than 12 days | 238 | 75.3 |
|  | 12 days and above | 78 | 24.7 |
| Time of admission | Day time | 247 | 78.2 |
|  | Night time | 69 | 21.8 |
| Type of surgery | Elective | 151 | 47.8 |
|  | Emergency | 110 | 34.8 |
| Amount of blood loss during procedure | <1500ml | 70 | 22.2 |
|  | ≥1500ml | 100 | 31.6 |
|  | Unrecorded | 146 | 46.2 |
| Type of anesthesia given during procedure | General anesthesia | 175 | 55.4 |
|  | Regional anesthesia (spinal or epidural) | 66 | 20.9 |
|  | Local anesthesia | 75 | 23.7 |
| Comorbid condition | Yes | 81 | 25.6 |
|  | No | 235 | 74.4 |
| Surgical site infection | Yes | 30 | 9.5 |
|  | No | 286 | 90.5 |
| Hospital acquired pneumonia | Yes | 26 | 8.2 |
|  | No | 290 | 91.8 |
| Medication used in the past 3 months | Yes | 15 | 4.7 |
|  | No | 301 | 95.3 |
| Medication used after admission at hospital | Yes | 316 | 100.0 |
|  | No | 0 | 0 |
| Total medication used after admission | ≤2 | 171 | 54.1 |
|  | >2 | 145 | 45.9 |

**Table 3:** Behavioral characteristics of the study participants at the surgical ward of ATRH and BDH, Arsi Zone, Oromia, Ethiopia, 2022 (n=316)

| Variables | Categories | Frequency | Percent |
| --- | --- | --- | --- |
| Body mass index | <18.5 kg/m<sup>2</sup> | 7 | 2.2 |
|  | 18.5-24.9 kg/m<sup>2</sup> | 296 | 93.7 |
|  | 25-29.9 kg/m<sup>2</sup> | 13 | 4.1 |
| Smoking status | Yes | 17 | 5.4 |
|  | No | 299 | 94.6 |
| Alcohol drinking status | Yes | 15 | 4.7 |
|  | No | 301 | 95.3 |

### 3.3 Behavioral characteristics of the study participants

From 316 of the study participants, 296(93.7%) had a BMI between 18.5 kg/m^2^ and 24.9 kg/m^2^ (see table three).

### 3.4 Factor associated with prolonged length of stay at surgical ward

In bivariate analysis factors that were candidate for multivariate analysis at p-value less than 0.2 were: Rural area, comorbid condition, regional anesthesia, blood loss ≥1500ml, unrecorded blood loss, regional anesthesia, surgical site infection, and hospital acquired pneumonia. However, in multivariable logistic regression analysis independent predictor of prolonged length of stay at p-value <0.05 were: Comorbid condition, surgical site infection, and hospital acquired pneumonia. The odd of prolonged length of stay among patients who had comorbid were 4.6 times [AOR=4.59, at 95% CI= (2.46-8.56)] more likely to have PLOS than who had no comorbid. The study participants who developed surgical site infection after surgery were 5 times [AOR=5.02 at 95% CI= 1.97-12.80)] more likely to experienced PLOS than their counterparts. The participants who developed hospital acquired pneumonia during their hospital stay were 3 times [AOR= 3.43 at 95% CI= (1.36-8.64)] more likely to experienced PLOS than those who had no pneumonia (see table 4).

**Table 4:** Bivariate and multivariate analysis of factor associated with prolonged length of stay among the study participants at surgical ward of ATRH and BDH, Arsi zone, Oromia, Ethiopia, 2022 (n=316)

| Variable | Categories | Prolonged length of stay |  | COR at 95% of CI | AOR at 95% of CI | p-value |
| --- | --- | --- | --- | --- | --- | --- |
|  |  | Yes | No |  |  |  |
|  |  | n<sub>o</sub>(%) | n<sub>o</sub>(%) |  |  |  |
| Place of residence | Rural | 46<br>(15%) | 117<br>(37%) | 1.49(0.89-2.49) | 1.45(0.79-2.65) | 0.225 |
|  | Urban | 32<br>(10%) | 121<br>(38%) | 1 | 1 |  |
| Comorbid condition | Yes | 39<br>(12.3%) | 42<br>(13.3%) | 4.67(2.68-8.13) | 4.59(2.46-8.56) | **0.000\*** |
|  | No | 39<br>(12.3%) | 196<br>(62%) | 1 | 1 |  |
| Amount of blood loss during surgery | <1500ml | 11<br>(3.5%) | 59<br>(18.7%) | 1 | 1 |  |
|  | ≥1500ml | 31<br>(10%) | 69<br>(22%) | 2.41(1.12-5.21) | 1.80(0.73-4.49) | 0.204 |
|  | Not recorded | 36<br>(11%) | 110<br>(34.8%) | 1.76(0.83-3.70) | 1.26(0.49-3.23) | 0.627 |
| Type of anesthesia | General anesthesia | 49<br>(15.5%) | 126<br>(40) | 1.15(0.62-2.12) | 0.36(0.11-1.16) | 0.086 |
|  | Regional anesthesia | 10<br>(3%) | 56<br>(17.7%) | 0.53(0.22-1.23) | 0.28(0.07-1.10) | 0.069 |
|  | Local anesthesia | 19<br>(6%) | 56<br>(17.7%) | 1 | 1 |  |
| Surgical site infection | Yes | 17<br>(5.4%) | 13<br>(4.1%) | 4.82(2.22-10.48) | 5.02(1.97-12.80) | **0.001\*** |
|  | No | 61<br>(19.3%) | 225<br>(71%) | 1 | 1 |  |
| Hospital acquired pneumonia | Yes | 13<br>(4.1%) | 13<br>(4.1%) | 3.46(1.53-7.83) | 3.43(1.36-8.64) | **0.009\*** |
|  | No | 225<br>(71.2%) | 65<br>(20.5%) | 1 | 1 |  |
| Total medication | ≤2 | 29<br>(9%) | 142<br>(45%) | 1 | 1 |  |
|  | >2 | 49<br>(15.5%) | 96<br>(30%) | 2.49(1.48-4.23) | 1.83(0.99-3.39) | 0.053 |

Note: \*p-value <0.05

## 6. Discussion

The study conducted among 316 patients revealed that the magnitude of PLOS was recorded in 78(24.7%) of the patients who stayed for at least 12 days based on the 75^th^ percentile cutoff point. The magnitude of PLOS of the current study is relatively lower than a study conducted at Oman tertiary hospital in Iran with PLOS rates of 30.5% (Almashrafi et al., 2016). This might be due to differences in study design and sample size. The magnitude of PLOS among adult patients admitted at SW of ATRH and BDH was in line with the study from Michigan University in America with PLOS rate of 22.4% (Krell et al., 2014), and the study conducted in JUMC with a PLOS rate of 25.3% (Tefera et al., 2020). But, the magnitude of current study is relatively higher than the study conducted in Mexico with PLOS of 5% (Marfil-Garza et al., 2018).The difference might be due to differences in sample size, and cutoff point to determine a PLOS which was 75^th^ percentile in the current study and 95^th^ percentile in Mexico.

This study finding indicated that patients with comorbid conditions were 4.6 times [AOR=4.59, at 95% CI= (2.46-8.56)] high likelihood of PLOS at the SW than those who had no comorbid. Similar to our study finding, a study conducted at Harvard University and American College of surgeon National Surgical Quality Improvement Program(ACSNSQI), 2009 revealed that patients with comorbid condition were experienced PLOS than their counterparts (Dasenbrock et al., 2015). The reasons why the patients with comorbidities experienced PLOS have not been fully evaluated. But, it might be related to that comorbid conditions delay the wound healing process by affecting blood transfusion toward the wound site and weakening the immune system that leading to extended duration of treatment which in turn increases hospital stay.

The current study showed that patients with surgical site infection were 5 times [AOR=5.02 at 95% CI= 1.97-12.80)] more likely to experienced PLOS than their counterparts. Similar to our study finding, a study conducted in German at the University of Hospital Magdeburg revealed that surgical site infection was one of the factors for PLOS (Jannasch et al., 2015). This might be due to patients with surgical site infection need additional management like medication and re-operation which lead to increase duration of treatment which in turn increase hospital stay.

This study also showed that patients who developed hospital acquired pneumonia during their hospital stay were 3 times [AOR= 3.43 at 95% CI= (1.36-8.64)] more likely to experienced PLOS than those who had no pneumonia. Similar study conducted in Harvard university revealed that patients with pneumonia were more likely to have PLOS (Dasenbrock et al., 2015).The reason for this is not fully evaluated but, it might be related to that it interrupt cutaneous wound healing through disruption of chemokine signals which is important for wound healing that in turn lead to increase duration of treatment and hospital stay.

### 6.1 Limitation of the study

Since consecutive sampling is nonprobability sampling, it lacks representativeness. Cross sectional study design cannot account for possible seasonal variation in disease incidence at SW and the study design was exposed to different type of bias. Lastly, one of the most common limitations in the current study was lack of literature in Ethiopia and Africa in the description of PLOS which limits the ability to compare and appropriately plan.

## 8. Conclusion and recommendation

The study finding showed that the magnitude of PLOS at SW is 24.7%. Surgical site infection, comorbid condition, and hospital acquired pneumonia were reported as the reason for experienced PLOS among the study participants. Based on the study finding quality of care could be improved by adjusting surgical ward environment to prevent hospital acquired infection and focus on managing complication after surgery. Health care provider should educate surgical patient about the risk of comorbidity on wound healing and early diagnosis and prevention of comorbid condition.

## Data Availability

All relevant data are within the manuscript and its Supporting Information files.

## Abbreviation

ATRH: Asella Teaching and Referral Hospital
BDH: Bekoji district hospital
BSC: Bachelor science BMI: Body mass index
IRB: Institutional review board
JUMC: Jimma university medical center
LOS: Length of stay
PLOS: Prolonged length of stay

## Data availability

Most of the data analyzed or used during this study are included in this published article, and additional files will be available from the corresponding author on reasonable request.

## Ethical approval

The ethical issue of this study was approved by the institutional review board (IRB) of Hawassa University.

## Consent

Respondents were informed about the objective and purpose of the study, and informed and verbal consent was obtained from each participants. The study participants were assured of the attainment of confidentiality for the information obtained from them.

## Conflict of interest

The author declare that they have no competing interest

## Authors’ Contributions

Deriba designed the study, perform analysis and interpretation of data, and drafted the manuscript. Gezahegn, Ezedin and Yohannis advised and supervised the design conception and made critical comments at each step of research. Jabir and Amdehiwot edited the manuscript. All authors read and approved the final manuscript. Confidentiality and anonymity were ensured throughout the execution of the study.

## Acknowledgments

First we would like to say thank to Madawalabu University for financial support. Next, the author would like to thank the study participants for their voluntary participation in the study.

## Notes

### Competing Interest Statement

The authors have declared no competing interest.

### Funding Statement

The author(s) received no specific funding for this work.

### Author Declarations

Ethical clearance was obtained from Hawassa University College of medicine and health science, the institutional review board (IRB)

